# Foundation model embeddings enable cardiovascular screening for people living with HIV in Vietnam using wearable signals

**DOI:** 10.1101/2025.10.11.25337800

**Authors:** Munib Mesinovic, Hai Ho Bich, Ly Vo Trieu, Viet Nguyen Quoc, Ngoc Nguyen Thanh, Tuan Anh Nguyen Hoang, Minh Tu Van Hoang, Phan Nguyen Quoc Khanh, Xuan Huy Vo, Phuc Vo Hong, Khoa Le Dinh Van, Yen Lam Minh, Louise Thwaites, Tingting Zhu

**Affiliations:** Department of Engineering Science, University of Oxford, Oxford, UK; Oxford University Clinical Research Unit, Ho Chi Minh City, Vietnam; Hospital for Tropical Diseases, Ho Chi Minh City, Vietnam; People’s Hospital 115, Ho Chi Minh City, Vietnam

**Keywords:** healthcare, HIV, interpretability, large language model, machine learning, time-series

## Abstract

Cardiovascular disease (CVD) screening faces significant challenges in resource-limited settings, where infrastructure and computational constraints preclude the use of advanced remote assessment. These constraints are particularly acute for people living with HIV (PLWH), who experience elevated CVD risk yet often receive care in clinics without the capacity for specialist diagnostics. We evaluate pretrained physiological embeddings from foundation models for CVD detection using low-cost wearable photoplethysmography (PPG) signals from 80 PLWH outpatients in Ho Chi Minh City, Vietnam. We compare a strictly zero-shot approach (NormWear applied without any local training) with a more practical pipeline that uses frozen PaPaGei embeddings plus a locally trained classifier. The PaPaGei-embedding approach achieved superior discrimination (AUROC 0.769) compared with zero-shot NormWear (0.610), traditional PCA features (0.651), and established clinical scores, including the Framingham score (0.551) and the D:A:D-modified Framingham score (0.462). Without fine-tuning the foundation model itself, PaPaGei embeddings captured clinically coherent structure: patients on dolutegravir-based regimens clustered in low-risk regions, while those with high cholesterol variability occupied high-risk areas, consistent with cardiometabolic pathophysiology. These results show that pretrained physiological embeddings can enable accurate screening when combined with lightweight local calibration, reducing reliance on extensive feature engineering while preserving pathophysiological plausibility and actionable triage behaviour in the learned embeddings. This provides a practical approach for deploying foundation models in resource-constrained settings where training deep models may be infeasible.

## 1 Introduction

Cardiovascular disease (CVD) screening is challenging to deliver at scale in resource-limited settings, where laboratory capacity, specialist diagnostics, and advanced computational infrastructure are often limited [1–3]. These structural barriers limit both traditional clinic-based approaches and data-hungry machine-learning pipelines that depend on large, annotated local datasets [4]. Practical, label-efficient strategies are therefore needed to enable robust triage within routine outpatient workflows. The challenge is particularly acute for people living with HIV (PLWH), who face a 1.5–2-fold higher risk of CVD than the general population [5–7]. In Vietnam, CVD accounts for 31% of all mortality, yet screening infrastructure remains limited and specialist services are concentrated in urban centres [8, 9]. These constraints hamper early identification of contemporaneous cardiac abnormalities among PLWH and complicate efforts to deploy conventional CVD detection tools or fully supervised models.

Despite the pressing need for scalable CVD screening in PLWH, machine learning approaches remain significantly underutilised in routine clinical practice for this population. A recent systematic review found that the vast majority of cardiovascular models for PLWH still rely on traditional statistical and simplistic linear approaches [10]. The limited adoption stems from multiple barriers: most ML models require extensive labelled datasets unavailable in resource-constrained settings [11], lack interpretability needed for clinical trust [12], and have not been validated in PLWH populations [13]. Furthermore, existing ML applications in cardiovascular medicine have predominantly focused on high-resource settings with advanced imaging modalities [14], leaving a critical gap for accessible, wearable-based approaches suitable for outpatient HIV care.

Photoplethysmography (PPG), obtainable via low-cost wearables, is an attractive modality for scalable, remote assessment [15, 16]. Heart rate variability (HRV) and waveform morphology features derived from PPG correlate with cardiovascular status and adverse events [17–19]. Pipelines based on handcrafted features and supervised classifiers, however, require careful preprocessing and disease-specific training data, limiting their generalisability across populations and care settings [20–23]. In PLWH specifically, established risk scores and conventional ML approaches have systematically underpredicted risk [24].

Foundation models trained on large-scale physiological data offer a practical alternative by providing rich pretrained representations that can be used with minimal local supervision. Models such as NormWear [25] and PaPaGei [26] extract embeddings from ECG and PPG that capture complex physiological structure. While these encoders can be applied in a strictly zero-shot manner, real-world deployment in low-resource clinics often benefits from lightweight local calibration—training only a simple classifier on frozen embeddings rather than fine-tuning the encoder—thereby balancing performance with computational feasibility.

Here, we evaluate pretrained physiological embeddings for CVD screening among 80 PLWH outpatients attending the clinic for routine medication collection in Ho Chi Minh City, Vietnam. We compare deployment modes ranging from strictly zero-shot application to a label-efficient pipeline using frozen PPG embeddings from PaPaGei with a locally calibrated classifier. We assess each encoder in its native modality (PaPaGei on PPG; NormWear on ECG/PPG for zero-shot baselines) to avoid crossdomain artefacts. We show that PaPaGei embeddings with a lightweight classifier achieve superior discrimination (AUROC 0.769) over zero-shot and clinical-score comparators, while aligning with known risk factors. This provides a practical framework for deploying foundation models in resource-constrained settings to support equitable, scalable CVD prescreening for PLWH.

## 2 Results

### 2.1 Dataset

We conducted a prospective study at the Hospital for Tropical Diseases in Ho Chi Minh City, Vietnam, enrolling 80 adults living with HIV (PLWH) between November 2023 and July 2024. All participants received routine antiretroviral therapy and had no acute illness at enrolment. Each participant was assessed for cardiovascular risk using Framingham and D:A:D-modified Framingham scores, and for evidence of cardiovascular disease using a standard 12-lead electrocardiogram (ECG) and echocardiography reviewed by expert cardiologists. In addition, wearable photoplethysmography (PPG) monitoring was performed using the SmartCare device. PPG waveforms were recorded at 100 Hz for approximately 20 minutes and then segmented into 5-minute windows. Conventional HRV and waveform morphology features were extracted for supervised baselines; our proposed approach uses short raw PPG segments with minimal preprocessing and no encoder fine-tuning. Follow-up visits occurred approximately every three months, with repeated PPG and ECG measurements.

We used binary labels indicating the presence or absence of cardiologist-confirmed CVD based on ECG and echocardiography interpretations. Participants were labelled CVD = 1 if clinically significant abnormalities were observed in either modality, and CVD = 0 otherwise. This binary label serves as ground truth for model evaluation; Framingham and D:A:D risk scores were retained as continuous comparators (see Methods). Of the 80 participants, 13 (16.3%) were classified as having cardiologist-confirmed CVD. Compared with CVD = 0, the CVD = 1 group was older (median 47 vs. 38 years), more frequently male (93% vs. 60%), and had higher systolic blood pressure (median 130 vs. 110 mmHg). Smoking, diabetes, and family history of CVD were also more prevalent in the CVD group. These features were used as inputs for supervised baselines, with ECG or echocardiographic evidence of CVD over 1 year serving as ground truth.

### 2.2 Baseline CVD Classification Using Clinical Features

We first evaluated the ability of supervised classifiers to detect ECG or echocardiographic evidence of CVD over 1 year using only static and summarised clinical variables collected over four outpatient visits, including age, sex, blood pressure, and cholesterol. Models were evaluated on a stratified held-out test set using 10 bootstrap resamples to estimate variability. As summarised in Table 2, Random Forest achieved the highest AUROC (0.744 *±* 0.08) and average precision (0.433 *±* 0.11), significantly outperforming the Framingham and D:A:D clinical risk scores (\*\**p <* 0.01**). However, all of the variables included in the Random Forest are also included in the traditional risk scores. Traditional risk scores showed limited discrimination for prevalent CVD in this cohort (Framingham AUROC 0.551; D:A:D 0.462). This is expected: the scores are designed to estimate future event risk, not to detect contemporaneous abnormalities. Here, we use them as continuous comparators for triage-style ranking; even under this generous interpretation, they underperformed the embedding-based pipeline.

**Table 1:**
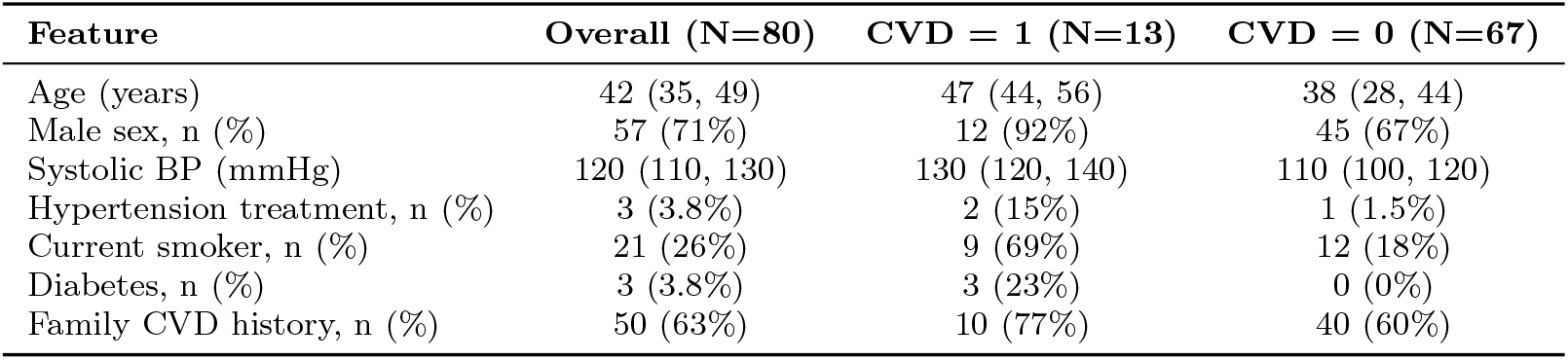
Baseline demographic and clinical features stratified by cardiologist-confirmed CVD status. Values are median (IQR) or n (%). Percentages are column-wise.

**Table 2:**
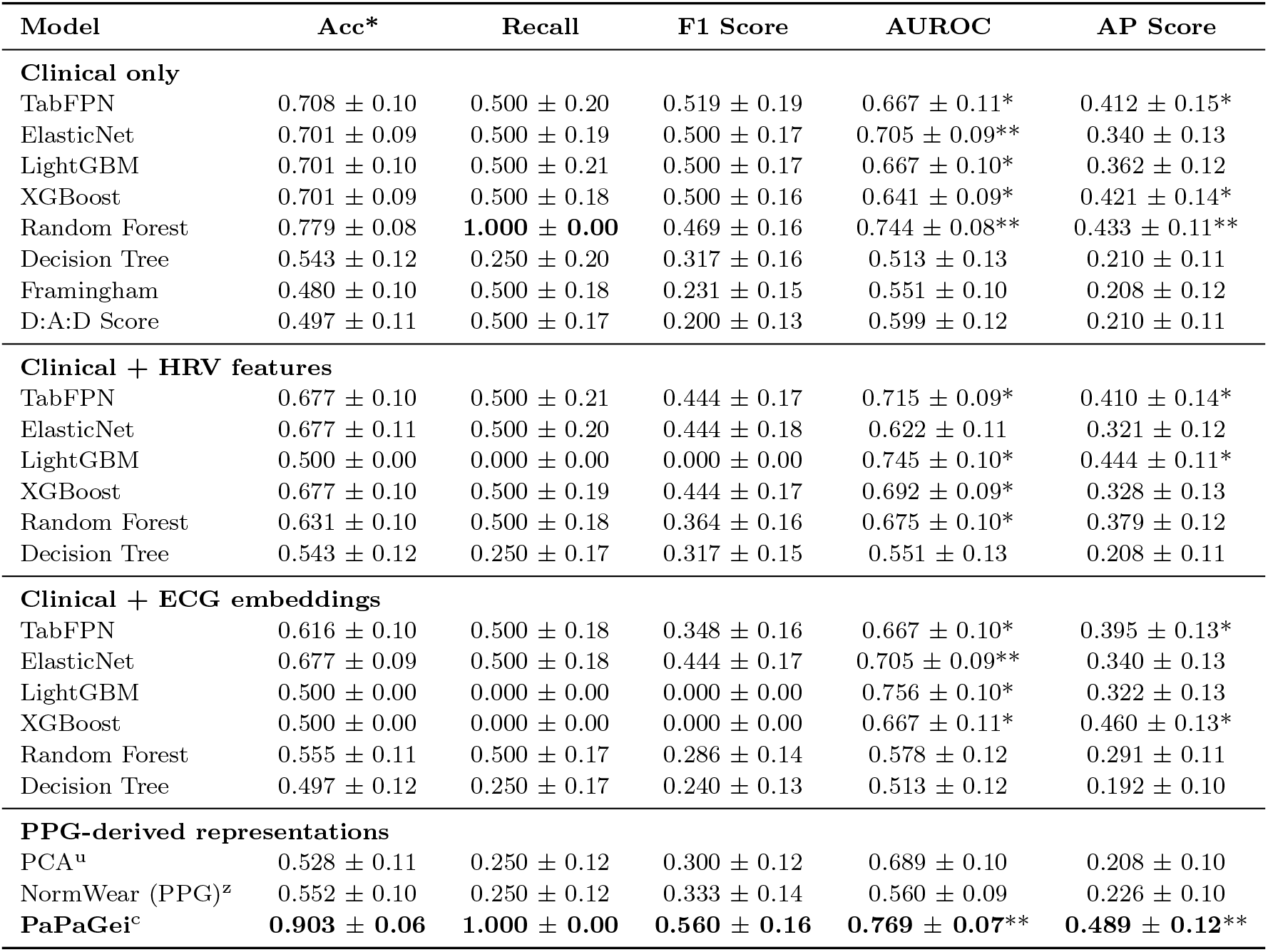
Performance of clinical, supervised, and zero-shot models for CVD classification. Models were trained or evaluated using clinical features, ECG embeddings, or raw PPG signals. NormWear was used in zero-shot mode; PaPaGei embeddings were evaluated using a downstream classifier. Metrics are reported as mean ± SD over 10 bootstrap resamples. D:A:D stands for D:A:D-modified Framingham score. For Framingham and D:A:D, AUROC/AP are computed on the continuous risk outputs as discrimination metrics against the prevalent CVD label; recall/F1/balanced accuracy use category-to-binary mappings (see Methods and Supplementary) and are provided only for operating-point context, not as assessments of 5–10-year calibration. Acc* stands for Balanced Accuracy. u = unsupervised; z = no local labels; c = frozen encoder with lightweight classifier. Bolded values indicate the best performance across all models. Asterisks indicate significant improvement over the Framingham score (\**p <*0.05, \*\**p <*0.01; Wilcoxon signed-rank test).

| Model | Acc* | Recall | F1 Score | AUROC | AP Score |
| --- | --- | --- | --- | --- | --- |
| **Clinical only** |  |  |  |  |  |
| TabFPN | 0.708 ± 0.10 | 0.500 ± 0.20 | 0.519 ± 0.19 | 0.667 ± 0.11* | 0.412 ± 0.15* |
| ElasticNet | 0.701 ± 0.09 | 0.500 ± 0.19 | 0.500 ± 0.17 | 0.705 ± 0.09** | 0.340 ± 0.13 |
| LightGBM | 0.701 ± 0.10 | 0.500 ± 0.21 | 0.500 ± 0.17 | 0.667 ± 0.10* | 0.362 ± 0.12 |
| XGBoost | 0.701 ± 0.09 | 0.500 ± 0.18 | 0.500 ± 0.16 | 0.641 ± 0.09* | 0.421 ± 0.14* |
| Random Forest | 0.779 ± 0.08 | **1.000 ± 0.00** | 0.469 ± 0.16 | 0.744 ± 0.08** | 0.433 ± 0.11** |
| Decision Tree | 0.543 ± 0.12 | 0.250 ± 0.20 | 0.317 ± 0.16 | 0.513 ± 0.13 | 0.210 ± 0.11 |
| Framingham | 0.480 ± 0.10 | 0.500 ± 0.18 | 0.231 ± 0.15 | 0.551 ± 0.10 | 0.208 ± 0.12 |
| D:A:D Score | 0.497 ± 0.11 | 0.500 ± 0.17 | 0.200 ± 0.13 | 0.599 ± 0.12 | 0.210 ± 0.11 |
| **Clinical + HRV features** |  |  |  |  |  |
| TabFPN | 0.677 ± 0.10 | 0.500 ± 0.21 | 0.444 ± 0.17 | 0.715 ± 0.09* | 0.410 ± 0.14* |
| ElasticNet | 0.677 ± 0.11 | 0.500 ± 0.20 | 0.444 ± 0.18 | 0.622 ± 0.11 | 0.321 ± 0.12 |
| LightGBM | 0.500 ± 0.00 | 0.000 ± 0.00 | 0.000 ± 0.00 | 0.745 ± 0.10* | 0.444 ± 0.11* |
| XGBoost | 0.677 ± 0.10 | 0.500 ± 0.19 | 0.444 ± 0.17 | 0.692 ± 0.09* | 0.328 ± 0.13 |
| Random Forest | 0.631 ± 0.10 | 0.500 ± 0.18 | 0.364 ± 0.16 | 0.675 ± 0.10* | 0.379 ± 0.12 |
| Decision Tree | 0.543 ± 0.12 | 0.250 ± 0.17 | 0.317 ± 0.15 | 0.551 ± 0.13 | 0.208 ± 0.11 |
| **Clinical + ECG embeddings** |  |  |  |  |  |
| TabFPN | 0.616 ± 0.10 | 0.500 ± 0.18 | 0.348 ± 0.16 | 0.667 ± 0.10* | 0.395 ± 0.13* |
| ElasticNet | 0.677 ± 0.09 | 0.500 ± 0.18 | 0.444 ± 0.17 | 0.705 ± 0.09** | 0.340 ± 0.13 |
| LightGBM | 0.500 ± 0.00 | 0.000 ± 0.00 | 0.000 ± 0.00 | 0.756 ± 0.10* | 0.322 ± 0.13 |
| XGBoost | 0.500 ± 0.00 | 0.000 ± 0.00 | 0.000 ± 0.00 | 0.667 ± 0.11* | 0.460 ± 0.13* |
| Random Forest | 0.555 ± 0.11 | 0.500 ± 0.17 | 0.286 ± 0.14 | 0.578 ± 0.12 | 0.291 ± 0.11 |
| Decision Tree | 0.497 ± 0.12 | 0.250 ± 0.17 | 0.240 ± 0.13 | 0.513 ± 0.12 | 0.192 ± 0.10 |
| **PPG-derived representations** |  |  |  |  |  |
| PCA<sup>u</sup> | 0.528 ± 0.11 | 0.250 ± 0.12 | 0.300 ± 0.12 | 0.689 ± 0.10 | 0.208 ± 0.10 |
| NormWear (PPG)<sup>z</sup> | 0.552 ± 0.10 | 0.250 ± 0.12 | 0.333 ± 0.14 | 0.560 ± 0.09 | 0.226 ± 0.10 |
| **PaPaGei<sup>c</sup>** | **0.903 ± 0.06** | **1.000 ± 0.00** | **0.560 ± 0.16** | **0.769 ± 0.07\*\*** | **0.489 ± 0.12\*\*** |

TabFPN yielded the highest precision (0.538 *±* 0.21) and F1 score (0.519 *±* 0.19), indicating strong discriminative ability. Random Forest, however, achieved perfect recall (1.000 *±* 0.00), maximising sensitivity to CVD cases at the cost of lower precision (0.306 *±* 0.17). In contrast, traditional risk scores performed poorly, with AUROCs of 0.551 (Framingham) and 0.462 (D:A:D), highlighting their limited applicability in this HIV-positive outpatient population. ElasticNet offered a good trade-off between discrimination and interpretability, with balanced metrics and an AUROC of 0.705 *±* 0.09. Overall, these results show that even in low-resource settings, simple supervised models trained on routine outpatient data can substantially improve CVD stratification, particularly compared to imported risk scoring systems.

### 2.3 Foundation Model Augmentation with ECG Embeddings

We next examined whether adding ECG-derived foundation-model representations to routine clinic data improves CVD prediction under label-limited conditions. Patient-level embeddings were extracted from raw 12-lead ECG using a frozen encoder (NormWear), reduced with PCA, and concatenated to static and temporal clinical features; no fine-tuning of the encoder was performed. NormWear primarily targets ECG but can produce zero-shot scores from PPG; we use it as a baseline without local training. This design tests whether a costlier signal (ECG) and a strong pretrained encoder improve deployable screening without fine-tuning in a small, imbalanced cohort of PLWH. PaPaGei is specialised for PPG and was not applied to ECG.

As summarised in Table 2, effects were model-dependent. ElasticNet remained stable relative to clinical-only inputs (Balanced Accuracy 0.677 ± 0.09, F1 0.444 ± 0.17). In contrast, tree ensembles frequently exhibited a small-*n* failure mode: LightGBM attained a high AUROC (0.756 ± 0.10) but defaulted to no positive calls (recall 0.000 ± 0.00; Balanced Accuracy 0.500 ± 0.00), with a similar pattern for XGBoost. The divergence between ranking (AUROC) and operating-point metrics (recall, Balanced Accuracy) indicates that while ECG embeddings contain a discriminative signal, a label-efficient alignment step is required to convert ranking into calibrated decisions at this scale.

These findings suggest limited incremental benefit from zero-fine-tuning ECG encoders over clinic data in our cohort, whereas the strongest gains arise in the PPG pathway when paired with a lightweight local calibrator.

### 2.4 CVD Detection Using Pretrained Embeddings with Lightweight Calibration

We evaluated whether pretrained physiological representations from wearable PPG can support cardiovascular disease detection (without any additional clinical data) under two deployment modes: a strictly zero-shot baseline, and a label-efficient pipeline with a frozen encoder and a lightweight local classifier. In the zero-shot setting, NormWear was applied without any local training to generate patient-level CVD probabilities from short PPG windows. As shown in Table 2, zero-shot performance was modest (AUROC 0.560, AP 0.226; Balanced Accuracy 0.552, F1 0.333), indicating that while some physiologically relevant signal is captured, discrimination is limited without local calibration. Including NormWear in a zero-fine-tuning PPG configuration provides the label-free reference point. The subsequent PaPaGei + calibrator pipeline is then taken as the minimal local adaptation required to unlock clinically usable performance.

In the label-efficient setting, PaPaGei was used as a frozen encoder to produce PPG embeddings, which were then passed to a calibrated Random Forest classifier trained on the local cohort. This pipeline yielded the highest discrimination among all methods evaluated (AUROC 0.769 ± 0.07, AP 0.489 ± 0.12), with strong operating characteristics (Balanced Accuracy 0.903 ± 0.06, recall 1.000 ± 0.00, F1 0.560 ± 0.16), and significantly outperformed the Framingham baseline (\*\**p <* 0.01**). Notably, this improvement was achieved without any fine-tuning of the foundation model itself.

As an unsupervised feature baseline, we applied principal component analysis (PCA) directly to raw PPG segments to obtain patient-level representations, followed by a Random Forest classifier. PCA retained some ranking ability (AUROC 0.689 ± 0.10), but class balance remained poor (Balanced Accuracy 0.528 ± 0.11, recall 0.250 ± 0.12; F1 0.300 ± 0.12), consistent with the low-signal, imbalanced setting and the absence of task-specific representation learning. Together, these results underscore the added value of pretrained PPG embeddings, with a minimal, on-site calibrator materially improving discrimination over both zero-shot scoring and unsupervised features.

To characterise how models stratify CVD, we visualised predicted probability distributions by ground-truth label (Figure 1). Framingham and D:A:D showed narrow, overlapping distributions, while a supervised Random Forest trained on clinical features improved separation primarily for low-risk patients. The PaPaGei + calibrator pipeline exhibited the clearest separation, concentrating high predicted likelihood of CVD almost exclusively among true CVD-positive cases, reflecting better calibration and sensitivity from raw PPG inputs.

**Fig. 1:**
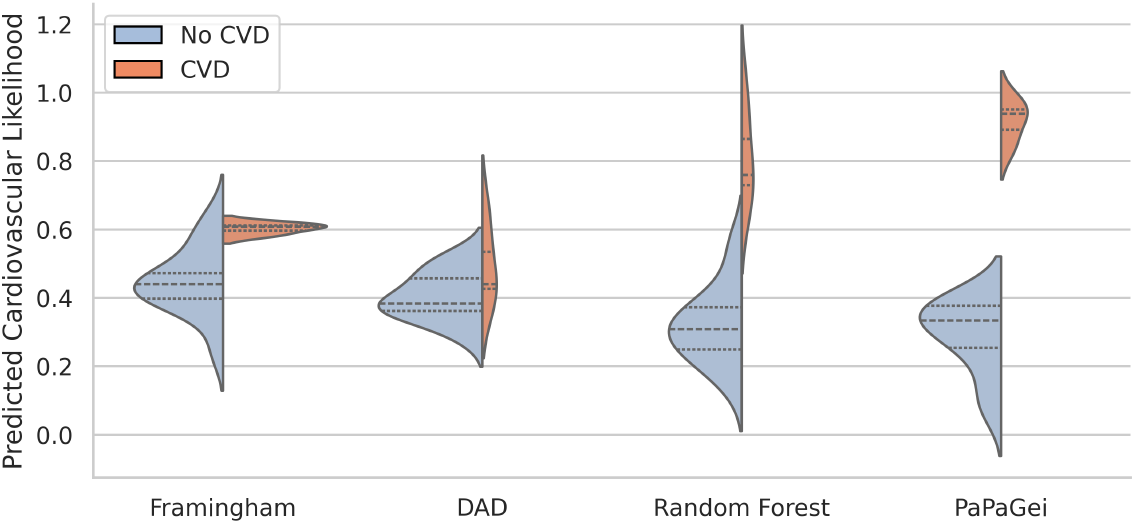
Predicted CVD likelihood distributions across models stratified by CVD label. We visualised predicted CVD probabilities for each model using violin plots split by ground-truth status (No CVD = blue, CVD = red). Traditional clinical risk scores (Framingham, D:A:D-modified Framingham) showed limited separation with overlapping distributions and compressed ranges. A supervised Random Forest trained on clinical features improved separation, particularly for low-risk individuals, but substantial overlap remained. In contrast, the PaPaGei + calibrator pipeline (frozen pretrained encoder with a lightweight local classifier) demonstrated a pronounced rightward shift for CVD-positive individuals, concentrating high predicted risk among true cases and indicating enhanced discriminative power from wearable PPG without fine-tuning the foundation model.

### 2.5 Embedding Separability and Feature Attribution

To better understand the structure of the learned patient representations, we visualised the embeddings produced by each model input type using three standard dimensionality reduction methods: PCA, UMAP, and t-SNE. As shown in Figure 2, patient-level features derived from clinical variables showed minimal class separability between individuals with and without cardiovascular disease (CVD). NormWear embeddings extracted from raw ECG signals provided marginal improvement, though the overlap between classes remained substantial. In contrast, PaPaGei embeddings computed directly from short PPG waveform segments demonstrated markedly clearer class separation across all projection methods. These findings suggest that PaPaGei captures richer, task-relevant physiological structure from raw PPG in a zero-shot setting, despite the absence of training on cardiovascular outcomes or ECG labels. PaPaGei has reported competitive performance on related PPG tasks, including cuffless blood pressure estimation and hypertension screening, suggesting its embeddings could capture haemodynamic structure relevant to our CVD-detection application [26]. This supports the potential utility of foundation model embeddings as standalone features for CVD probability stratification, particularly when traditional clinical features or labelled training data are limited.

**Fig. 2:**
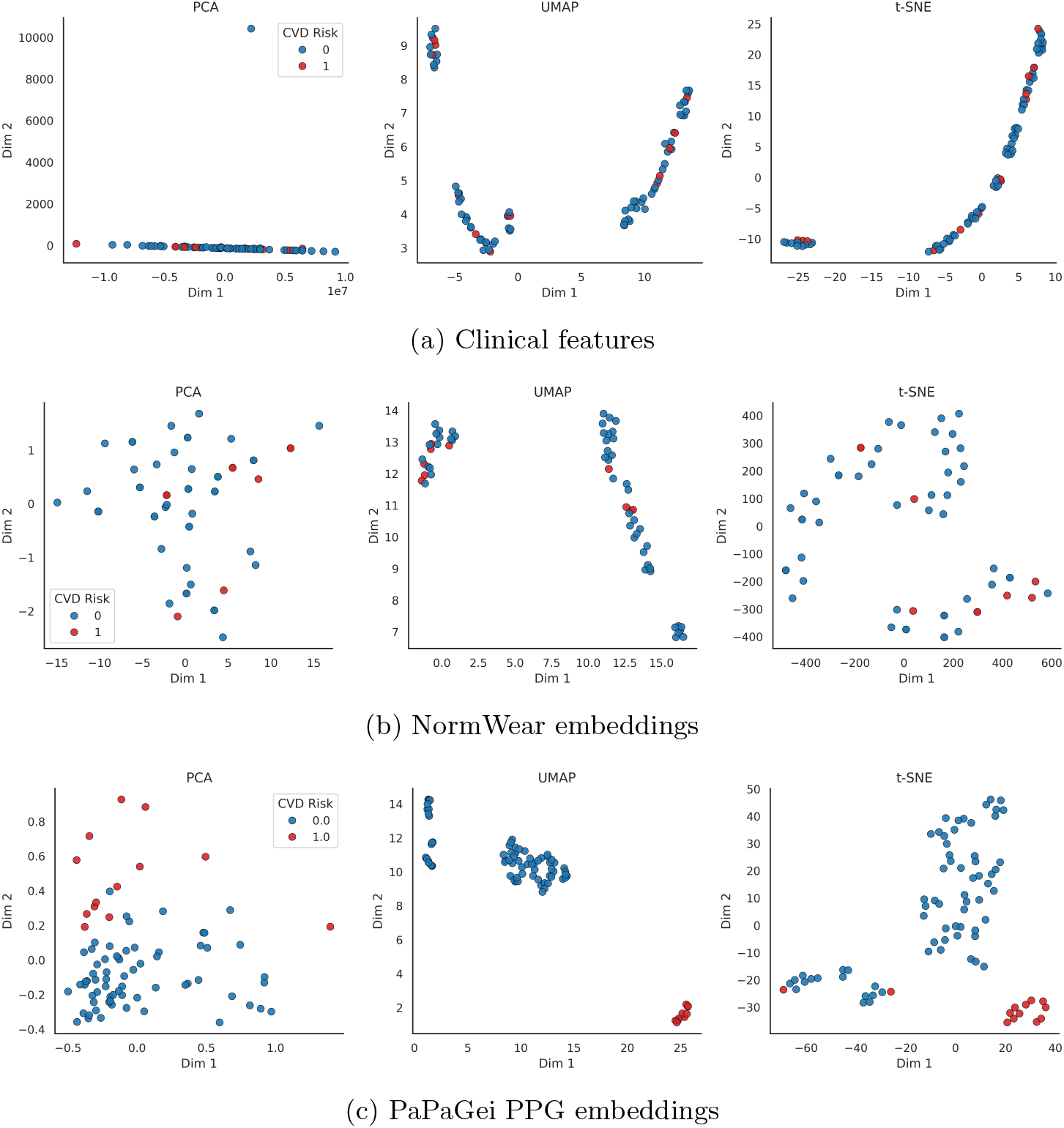
Latent space projections of patient representations by model input type. We visualised the separability of patient-level CVD likelihood using PCA, UMAP, and t-SNE projections on (a) routine clinical features, (b) NormWear embeddings, and (c) PaPaGei PPG embeddings. Compared to clinical and ECG-derived representations, PaPaGei embeddings showed markedly better class separation between CVD and non-CVD patients across all dimensionality reduction methods. Compared to clinical and ECG-derived representations, PaPaGei embeddings showed markedly better class separation between CVD and non-CVD participants across all dimensionality reduction methods. This suggests that PaPaGei captures richer physiological signals relevant to cardiovascular status from raw PPG with a frozen encoder (no fine-tuning).

To further investigate whether PaPaGei embeddings encode clinically meaningful signals, we overlaid selected clinical variables, previously identified as important by supervised models, onto the projected embedding space (Figure 3). Notably, patients receiving the antiretroviral regimen Tenofovir/lamivudine/dolutegravir (TDF 3TC DTG) appeared largely confined to low-risk regions of the latent space, supporting its observed protective association with CVD absence. Conversely, larger changes in total cholesterol across visits (TOTAL CHOL delta), a known cardiovascular risk marker, were concentrated in high-risk clusters where CVD-positive patients were more prevalent. These overlays reveal alignment between PaPaGei’s unsupervised physiological representations and clinically validated risk factors, reinforcing the model’s capacity to capture latent structure that is both discriminative and biologically informative. This interpretability also demonstrates how foundation models may serve as hypothesis-generating tools, highlighting patterns of treatment response or physiological dysregulation that merit further investigation.

**Fig. 3:**
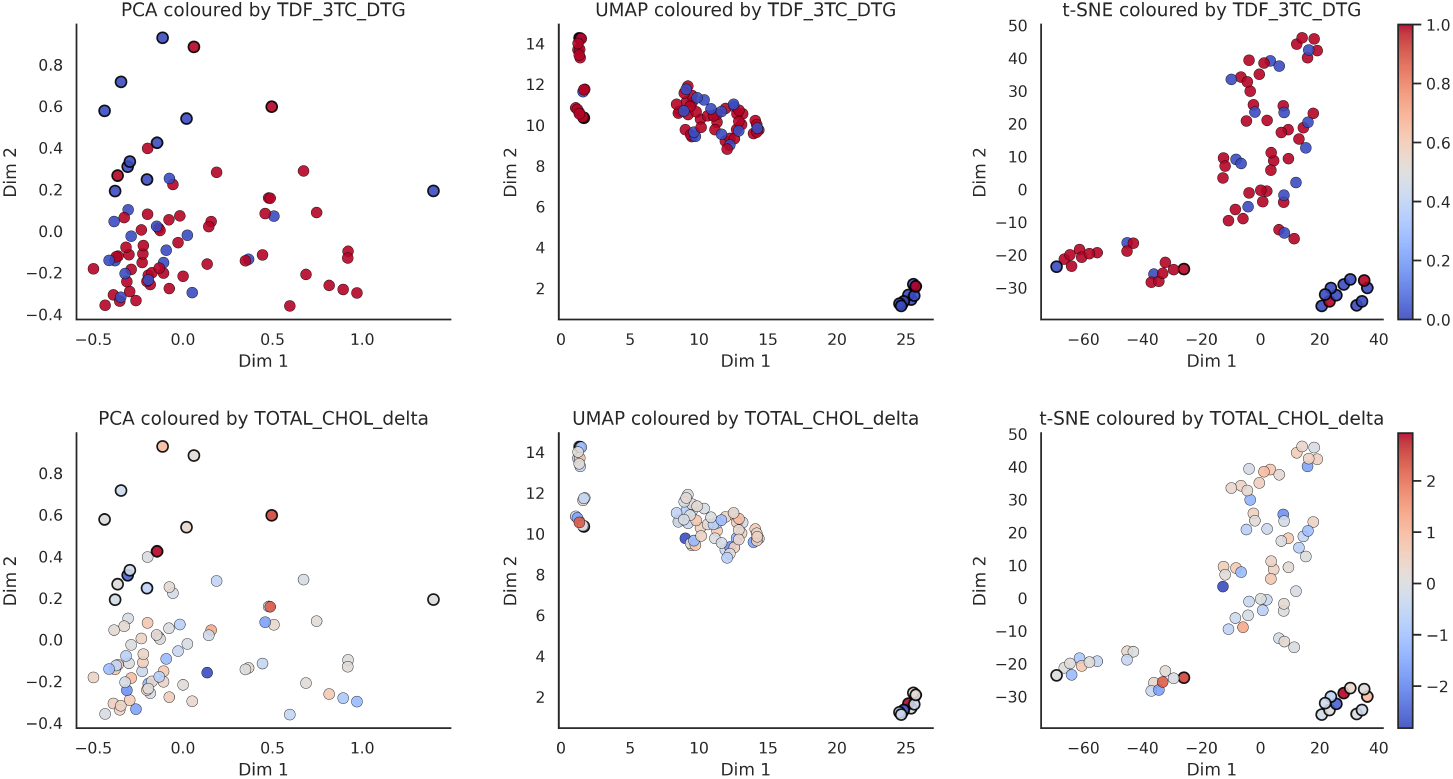
Clinical variable overlays on PaPaGei PPG embedding space. We visualised the projections of patient-level PaPaGei embeddings using PCA, UMAP, and t-SNE, overlaid with clinically relevant variables. Dot outlines indicate binary cardiovascular disease (CVD) status (black = CVD, white = no CVD), while colour represents values of selected clinical predictors. (**Top row**) TDF 3TC DTG usage is associated with a lower density of CVD-positive patients, consistent with protective antiretroviral therapy effects. (**Bottom row**) Changes in total cholesterol (TOTAL CHOL delta) across visits are higher in the CVD-positive clusters, supporting its predictive relevance. These overlays confirm that clinical variables identified as important by supervised models align with separable physiological patterns learned by PaPaGei without encoder fine-tuning.

## 3 Discussion

This study demonstrates that pretrained physiological embeddings from foundation models can enable accurate cardiovascular screening when combined with lightweight local calibration. We evaluated two pragmatic scenarios: strictly zero-shot deployment (NormWear) and a hybrid approach using frozen PaPaGei embeddings with a locally-trained classifier. The PaPaGei pipeline achieved superior performance (AUROC 0.769), substantially outperforming both the zero-shot baseline and traditional risk scores. Traditional risk scores and our PPG-based approach target different clinical endpoints: population-level prediction of future events versus contemporaneous prescreening for abnormalities warranting confirmatory testing. While Framingham and D:A:D-modified Framingham scores appropriately predict future events for population-level prevention strategies, our method identifies current cardiovascular abnormalities requiring immediate clinical attention.

Importantly, while our approach required local labels to train the classifier, it avoided the computational burden of fine-tuning the foundation model itself. The PaPaGei embeddings remained frozen throughout, preserving their pretrained representations while a simple Random Forest learned to map these features to cardiovascular disease likelihood. PaPaGei has also been somewhat validated for bias across skin tones with raw PPG signal data and has shown robust performance. This strategy is particularly relevant for resource-limited settings where deep learning infrastructure is unavailable but basic machine learning tools remain feasible.

The clinical plausibility of the learnt latent structure further supports deployment. PaPaGei embeddings organised patients on dolutegravir-based regimens into low-risk regions, aligning with evidence for cardiometabolic benefits of newer antiretrovirals [27–29]. Conversely, greater visit-to-visit cholesterol variability concentrated in high-risk regions, consistent with known associations between lipid instability and cardiovascular events in HIV [30, 31]. These patterns emerged without any HIV-specific fine-tuning, indicating that pretrained representations capture physiology that transfers across populations and care contexts.

From an implementation perspective, this embedding-based approach offers a practical middle ground. While not truly “zero-shot,” it dramatically reduces the complexity compared to training deep models from scratch. The foundation model handles feature extraction, requiring only a lightweight classifier to be trained locally. This could be accomplished with standard computing resources available in most clinical settings, unlike the GPU infrastructure needed for deep learning. We emphasise that PPG is not a clinician-interpreted signal; the utility here derives from embeddings that encode physiological structure and support triage decisions when paired with a shallow calibrator.

Our single-site cohort is small and class-imbalanced, however. While bootstrap resampling provides uncertainty estimates, larger multi-site evaluations are needed to characterise stability across devices, acquisition conditions, and case mix [32]. Although PaPaGei was never fine-tuned on local outcomes, the use of a shallow calibrator means performance reflects a label-efficient rather than strictly label-free configuration. Finally, potential spectrum and selection biases, as well as choices in signal preprocessing, windowing, and embedding aggregation, may influence generalisability. Our comparison between current disease detection and likelihood (foundation models) and future risk prediction (clinical scores) represents different clinical endpoints.

While this study was not a formal economic evaluation, we outline pragmatic considerations for deploying a wearable PPG + foundation-model pipeline as a prescreening tool in HIV outpatient care. Echocardiography (and clinician-interpreted ECG) provides structural and electrophysiological assessment that PPG cannot. Our approach is intended for triage, identifying PLWH who may warrant confirmatory testing, not for replacing imaging. In this role, the appropriate comparators are clinic-visit risk scores and simple supervised models using routinely collected variables, for which our embedding + lightweight-calibrator pipeline showed superior discrimination in this cohort. Short PPG captures can be obtained in the waiting area by non-specialist staff or remotely via wearables and scored on commodity hardware (frozen encoder; shallow classifier), reducing dependence on laboratory testing and specialist time within a single outpatient encounter. A formal micro-costing study is an important next step to quantify these trade-offs.

Future work should prioritise prospective, multi-centre validation in Vietnam, calibration and threshold selection for operational triage, and cost-effectiveness modelling to balance screening coverage against referral capacity. Methodologically, we find two directions to be most promising: learning-curve analyses to quantify label efficiency (how performance scales from a handful of labels), and strictly labelfree anomaly scoring over foundation-model embeddings (e.g., mixture modelling or distance-to-normative embeddings) as a complement to the lightweight calibrator. Fairness analyses across sex, age, and antiretroviral regimens are also important for real-world deployment.

In conclusion, pretrained physiological models enable low-cost cardiovascular screening without task-specific fine-tuning and with only minimal local supervision, outperforming conventional risk scores that underperform in HIV populations [24, 33]. Together with a strictly zero-shot baseline that delivers disease discrimination, these findings outline a practical path for deploying AI-driven screening where labelled data and specialist resources are limited, supporting more equitable access to cardiovascular care for people living with HIV.

## 4 Methods

### 4.1 Data acquisition and processing

This study was conducted at the Hospital for Tropical Diseases in Ho Chi Minh City, Vietnam, between 17 November 2023 and 20 July 2024. The protocol was approved by the institutional ethics committee and the Oxford Tropical Research Ethics Committee, with all participants providing written informed consent. Eligible individuals were adults (*≥*18 years) receiving routine outpatient HIV care with no acute illness or indication for hospital admission. Further details can be found in Supplementary Section 1.

At baseline, each participant completed a standardised assessment including demographic variables (age, sex, year of HIV diagnosis, smoking history), antiretroviral therapy (ART) regimen, medication use, systolic and diastolic blood pressure, and anthropometric measures. Laboratory tests included total cholesterol, HDL cholesterol, CD4 count, and HIV viral load. Cardiovascular risk was assessed using both the Framingham score and its D:A:D-modified version, each calculated using baseline clinical and laboratory inputs. Participants also underwent 12-lead electrocardiography (ECG), transthoracic echocardiography, and wearable photoplethysmography (PPG) monitoring using a SmartCare pulse oximeter (SmartCare Analytics UK). ECG and echocardiogram interpretations were independently reviewed by two cardiologists, with CVD-positive status assigned if either modality showed clinically significant abnormalities warranting further investigation or treatment.

Follow-up visits occurred every 3 months and included repeat ECG, PPG, and cardiovascular risk scoring. PPG signals were acquired using the SmartCare BM2000A wearable pulse oximeter following a standardised protocol (see Supplementary Section 4 and Figure S1 for device setup details). The non-invasive monitoring setup enabled data collection during routine clinic visits without specialised infrastructure. PPG waveforms were recorded at 100 Hz via Bluetooth transmission to a mobile application (SmartCare Capture, available on Google Play Store). Recordings lasted approximately 20 minutes, with the first 15 minutes used for analysis. Raw PPG signals underwent standardised preprocessing and quality control using the Vital SQI package [34], including noise filtering and segment rejection. Handcrafted heart rate variability (HRV) and waveform morphology features were extracted using the Vital DSP [35] library. HRV features were summarised across all segments per participant by computing the mean, standard deviation, minimum, and maximum of each feature.

Clinical features were derived from a combination of single-timepoint variables (Visit 1) and multi-visit time series. Time-varying features, systolic/diastolic blood pressure and cholesterol, were collected across four outpatient visits and imputed row-wise using each patient’s mean value where missing. Temporal statistics (mean, standard deviation, min, max, range, slope, and delta between Visit 1 and Visit 4) were computed per feature to capture dynamic risk patterns. ART regimen variables, smoking status, and cardiovascular history were retained as binary indicators. Derived features such as years on ART (year of treatment minus diagnosis) and categorical medication usage were also included. The Framingham and D:A:D-modified Framingham risk scores were discretised to binary indicators for comparison. For Framingham, moderate and high risk categories (score = 2) were mapped to 1, and low risk to 0. For D:A:D, we mapped high and very high risk categories (scores 2 and 3) to 1.

All supervised machine learning models were implemented in Python using scikit-learn and https://github.com/PriorLabs/TabPFN. We trained Random Forest, ElasticNet, LightGBM, XGBoost, TabFPN, and Decision Tree classifiers using demographic and temporal clinical variables, with or without HRV features. For ElasticNet, the regularisation parameters were tuned using Bayesian optimisation; for tree-based models, we used fixed or lightly tuned hyperparameters to reflect practical clinical deployment. Training details can be found in Section 2 of the Supplementary Material. Foundation model embeddings were extracted using PaPaGei for PPG signals and NormWear for ECG, each in zero-shot mode without downstream fine-tuning. PaPaGei embeddings were reduced using PCA and passed to a calibrated Random Forest classifier. NormWear outputs were averaged across segments per patient. All comparisons used expert-verified CVD status as ground truth.

### 4.2 Model Implementation

To evaluate CVD detection from physiological time series under both zero-shot and label-efficient regimes, we employed two foundation models: NormWear [25] and PaPaGei [26]. NormWear was applied to the raw ECG to extract embeddings for augmenting clinical features. For PPG, we evaluated a strictly zero-shot baseline using NormWear applied without any local training to produce patient-level probabilities, and a label-efficient pipeline using PaPaGei as a frozen encoder to produce embeddings that were fed to a lightweight calibrated Random Forest classifier. NormWear is included for two reasons: as a modality-matched comparator for ECG when augmenting clinic data without fine-tuning, and as a zero-fine-tuning PPG baseline that reflects the most label- and compute-constrained deployment scenario. This prevents cherry-picking and defines a lower bound that more practical, label-efficient pipelines must exceed. PCA on raw PPG served as an unsupervised feature baseline.

In contrast, PaPaGei was used to extract general-purpose embeddings from short PPG segments in a frozen-encoder setting. An overview of this pipeline is shown in Figure 4. Raw PPG waveforms sampled at 100 Hz were segmented into overlapping 10-second windows (stride = 5 seconds). Each window underwent preprocessing, including denoising, detrending, z-score normalisation, and signal quality filtering. Only windows passing noise and flatline thresholds were retained. These were then passed to the PaPaGei foundation model via prompt-based inference to produce fixed-length physiological embeddings. Prompt templates used are detailed in the Supplementary Materials section 3.

**Fig. 4:**
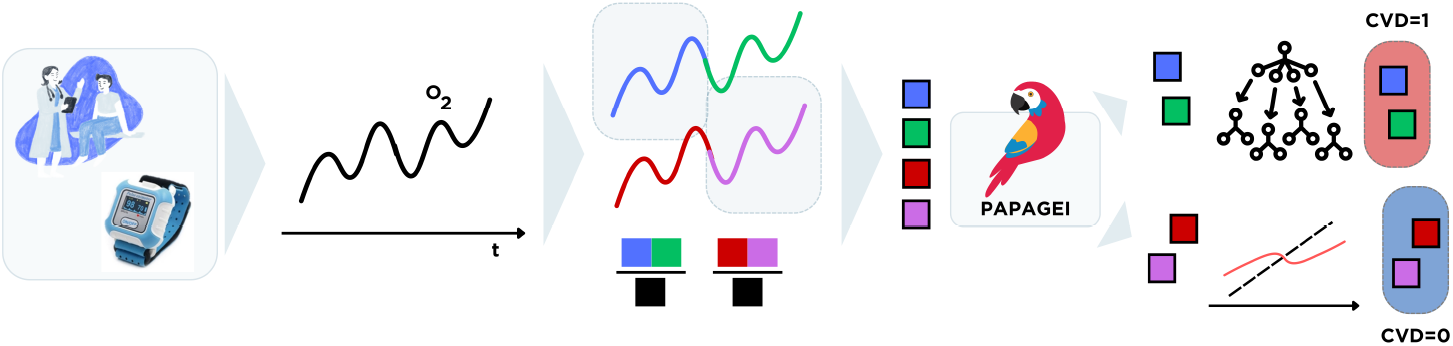
End-to-end embedding-based screening pipeline using PaPaGei. PPG signals were recorded from PLWH using low-cost wearable pulse oximeters (SmartCare) during clinic visits. Signals were segmented into 10-second windows, quality filtered, and normalised before being passed through the PaPaGei foundation model using structured prompting. The resulting physiological embeddings were aggregated at the patient level and passed to a calibrated Random Forest classifier. This pipeline achieved the highest predictive performance across all tested configurations (AUROC = 0.769, AP = 0.489, recall = 1.000), outperforming traditional clinical risk scores and supervised baselines. The approach requires no encoder fine-tuning and no laboratory data, and uses a lightweight local classifier for calibration, making it viable for scalable prescreening in outpatient workflows.

For each patient, embeddings across windows were mean-pooled and reduced via principal component analysis (PCA, retaining 15 components) to form a compact patient-level representation. These representations were input to a Random Forest classifier trained to predict CVD status. Classifier calibration was performed using isotonic regression on the training set. Evaluation was conducted on a stratified test set across 10 bootstrap iterations. Performance metrics, including AUROC, average precision, recall, precision, and F1 score, are reported in Table 2.

To interpret the learned representations, we projected the PCA-reduced embeddings using PCA, t-SNE, and UMAP, and visualised them with respect to ground truth labels and clinically important covariates. As shown in Figures 2 and 3, PaPaGei embeddings exhibited strong class separability and aligned with key clinical predictors such as antiretroviral regimen and lipid profile changes. These results suggest that PaPaGei captures physiologically relevant structure from wearable PPG in a zero-shot setting.

### 4.3 Evaluation

All models were trained and evaluated using stratified 80/20 train-test splits to preserve the proportion of cardiovascular disease (CVD) cases in both sets. To assess model stability and account for small sample variation, we generated 10 bootstrap resamples of the test set. Model performance was reported as the mean and standard deviation of metrics across these resamples.

For models with tunable hyperparameters, including ElasticNet and tree-based methods, we conducted internal validation using stratified 3-fold cross-validation on the training split. ElasticNet hyperparameters (penalty and mixing ratio) were optimised via Bayesian optimisation to maximise the area under the receiver operating characteristic curve (AUROC). This approach adaptively selects hyperparameter combinations to explore based on surrogate modelling of performance, offering efficiency over grid or random search.

We evaluated model discrimination using the AUROC and average precision (AP), and binary classification performance using precision, recall, and F1 score. Let *ŷ*_*i*_ *∈ {*0, 1*}* denote the predicted label and *y*_*i*_ *∈ {*0, 1*}* the ground truth for sample *i*. Then:

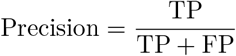

where TP is the number of true positives and FP is the number of false positives.

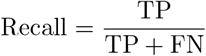

where FN is the number of false negatives.

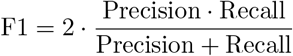

**AUROC** (Area Under the Receiver Operating Characteristic Curve) quantifies the ability to rank positive instances above negative ones across all classification thresholds. It is equivalent to the probability that a randomly chosen positive instance has a higher predicted score than a randomly chosen negative one.

**Average Precision (AP)** summarises the precision-recall curve by computing the weighted mean of precisions achieved at each threshold:

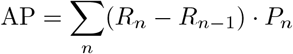

where *P*_*n*_ and *R*_*n*_ are the precision and recall at threshold *n*.

For statistical comparison of model performance, we conducted pairwise Wilcoxon signed-rank tests on AUROC and AP scores across the 10 test resamples. This non-parametric test accounts for paired, non-normally distributed data and is appropriate for small-sample comparative evaluation. Statistical significance was defined as *p <* 0.05, with results annotated accordingly in Table 2.

## Data Availability

Patient data and individual-level source data in this article cannot be made publicly available without further consent and ethical approval owing to privacy concerns.

## Code availability

Code for running and analysing the models can be found here: https://github.com/munibmesinovic/CVD-HIV.git.

## Acknowledgements

MM is supported by the Rhodes Trust and the EPSRC CDT Health Data Science. TZ is supported by the Royal Academy of Engineering.

## Author contributions

## Supplementary information

Supplementary material is contained in a separate file.

