## Supplementary Materials for "Foundation model embeddings enable cardiovascular screening for people living with HIV in Vietnam using wearable signals"

### 1 Data

#### 1.1 Study Population and Setting

This study enrolled 80 asymptomatic adults living with HIV (PLWH) attending the outpatient department of the Hospital for Tropical Diseases (HTD) in Ho Chi Minh City, Vietnam, between 17th November 2023 and 20th July 2024. Eligibility criteria included age ≥18 years and stable HIV treatment. Patients with acute illness or current hospitalisation were excluded. Written informed consent was obtained from all participants. The study was approved by the HTD Ethics Committee and the Oxford Tropical Research Ethics Committee.

Participants were followed up over four outpatient visits across a 12-month period. All 80 patients attended Visit 2, 79 attended Visit 3, and 74 attended Visit 4. At each visit, participants underwent wearable monitoring, ECG, and a brief acceptability survey.

#### 1.2 Cardiovascular Risk Assessment and Diagnosis

Baseline risk was assessed using the Framingham 10-year cardiovascular risk score, modified using the D:A:D HIV-specific criteria, which incorporate additional antiretroviral therapy exposure features. Patients were stratified into "low" or "higher" cardiovascular risk groups. Conventional 12-lead ECG and echocardiography were performed at baseline, with ECG repeated at follow-up visits. ECG and echo findings were independently reviewed by two cardiologists to identify individuals with abnormalities warranting further cardiovascular evaluation. Patients with either ECG or echocardiographic abnormalities were considered CVD-positive. A total of 13 out of 80 participants (16.3%) met this criterion during the study.

#### 1.3 Wearable Signal Acquisition and Feature Extraction

PPG waveforms were collected using SmartCare wearable pulse oximeters (SmartCare Analytics UK), transmitting data via Bluetooth at 100 Hz to an Android mobile application ("SmartCare Capture"). Each participant was monitored for approximately 20 minutes. After quality control (noise and flatline detection), over 95% of signal data were retained. From these recordings, five-minute high-quality PPG segments were selected for each patient and used for physiological feature extraction.

Figure 1 shows the complete SmartCare BM2000A pulse oximeter setup used for PPG data acquisition. The device kit included: (1) SmartCare watch with OLED display, (2) adjustable wrist strap, (3) finger sensor cable with red/infrared LEDs, (4) USB charging cable, and (5) Samsung tablet with SmartCare Capture application for real-time visualisation and data storage.

Participants were seated comfortably in the outpatient waiting area. The following standardised protocol was followed:

1. Attach the finger sensor cable to the SmartCare watch main unit

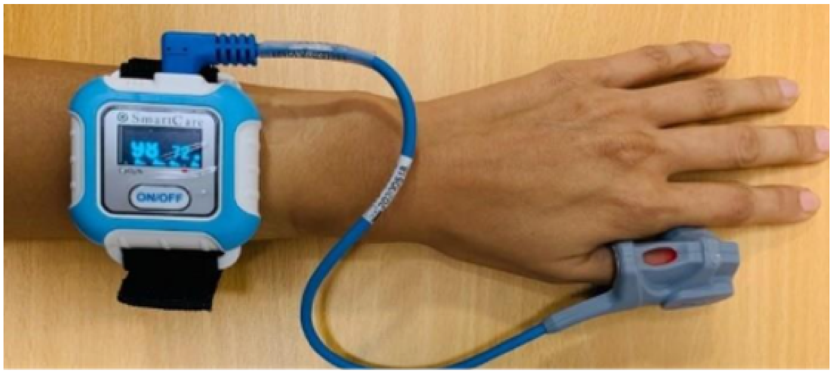


Figure 1: SmartCare wearable pulse oximeter components and patient setup. (a) Complete device kit showing the wrist unit, finger sensor, and data collection tablet. (b) Proper sensor placement with nail-side orientation for optimal signal quality. (c) Real-time PPG waveform display on the SmartCare Capture application during a 20-minute recording session. The CE-marked device transmitted data via Bluetooth at 100 Hz, with quality control achieving >95% usable signal retention across all participants.

2. Secure the device to the participant's non-dominant wrist using the adjustable strap
3. Position the finger sensor on the index or middle finger with the nail symbol facing upward
4. Power on the device and verify red sensor light activation
5. Confirm Bluetooth connection to the paired tablet (within 5-10 meters)
6. Monitor real-time signal quality via SmartCare Capture application
7. Record continuously for 20 minutes, with the first 15 minutes used for analysis
8. After completion, disconnect via the app before removing the device

Signal quality was monitored throughout by trained clinic staff. Participants were instructed to remain seated with minimal arm movement during recording. The device automatically filtered motion artifacts and provided real-time feedback on signal quality through the tablet interface.

Heart rate variability (HRV) and PPG morphology features were extracted using the open-source `vital_DSP` and `vital_SQI` Python packages. In total, 61 HRV and waveform morphology features were computed per patient. Selected features include time-domain (e.g., RMSSD, SDNN), frequency-domain (e.g., LF/HF power), non-linear (e.g., Poincaré, DFA), and morphological (e.g., systolic slope, diastolic duration) metrics.

##### 1.4 Clinical Feature Compilation

Baseline clinical and demographic variables were collected, including age, sex, treatment history, smoking status, and systolic/diastolic blood pressure. Additional features were derived from up to four outpatient visits, including total cholesterol, HDL, and systolic blood pressure. Time-varying features were aggregated via mean, delta, slope, min/max, and range. A full list of clinical features used for modelling is summarised in Table 1.

##### 1.5 Cardiovascular Disease Prevalence and Risk Score Distributions

Of the 80 patients, 13 were found to have ECG or echocardiographic evidence of CVD. Risk score distributions were skewed: 65% (52/80) were classified as low-risk by Framingham, compared to 45% (36/80) by D:A:D. Importantly, several CVD-positive patients were misclassified as low-risk by both scores, highlighting the limitations of existing screening tools in this population.

##### 1.6 Summary of Patient Characteristics and Feature Distributions

Table 1 summarises clinical, demographic, and waveform-derived feature distributions across the full cohort of 80 participants. Continuous variables are reported as median (IQR); categorical variables are reported as $n$ (%).

#### 2 Training Details

##### 2.1 Model Implementation and Frameworks

All supervised machine learning models were implemented in Python using `scikit-learn` (v1.3.0) and related packages including `xgboost`, `lightgbm`, and `tabpfn`. Model development, hyperparameter tuning, and evaluation were conducted using reproducible pipelines with fixed seeds (42) for all random splits.

All preprocessing steps were performed using `pandas`, `numpy`, and `scikit-learn` preprocessing modules. Data was split into stratified training and held-out test sets (80/20), with bootstrapping used to estimate performance variability across 10 resamples. Model calibration was conducted using isotonic regression fit on the training set. Experiments were conducted on a MacBook Pro (Apple M3 Pro chip, 51 GB RAM) without GPU acceleration, reflecting the resource-constrained, low-cost settings in which our approach is designed to operate.

##### 2.2 Supervised Model Training and Tuning

The following models were implemented and evaluated:

Table 1: Baseline and waveform feature distributions for all 80 participants. Values are shown as median (IQR) or count (%).

| **Feature** | **Value (N = 80)** |
|---|---|
| **Demographics** | |
| Age (years) | 42 (35, 49) |
| Sex (Male) | 57 (71%) |
| Current smoker | 21 (26%) |
| Diabetes | 3 (3.8%) |
| Hypertension (treatment) | 5 (6.3%) |
| Family history of CVD | 50 (63%) |
| **Vital Signs and Labs** | |
| Systolic BP (mmHg) | 120 (110, 130) |
| HDL Cholesterol (mmol/L) | 1.09 (0.94, 1.25) |
| Total Cholesterol (mmol/L) | 4.29 (3.83, 4.79) |
| CD4 Cell Count (cells/$mm^3$) | 510 (391, 623) |
| **Framingham Score (10yr)** | |
| Low risk | 52 (65%) |
| High risk | 28 (35%) |
| **D:A:D Modified Score** | |
| Low risk | 36 (45%) |
| High risk | 44 (55%) |
| **HRV Features (PPG-derived)** | |
| SDNN (ms) | 155 (71, 304) |
| RMSSD (ms) | 210 (93, 431) |
| pNN50 (%) | 13 (5, 32) |
| Mean NN (ms) | 818 (746, 921) |
| Total Power ($ms^2$) | 6,032 (1,496, 16,578) |
| LF/HF Ratio | 0.69 (0.52, 1.00) |
| DFA | 0.90 (0.84, 0.98) |
| **PPG Morphology Features** | |
| Systolic duration (s) | 0.43 (0.40, 0.47) |
| Diastolic duration (s) | 0.39 (0.34, 0.45) |
| Systolic slope | 1.55 (1.44, 1.56) |
| Diastolic slope | -1.563 (-1.568, -1.553) |
| Systolic amplitude variability | 6,021 (4,776, 6,970) |
| Heart rate (bpm) | 76 (68, 84) |

- Logistic Regression with ElasticNet regularisation (`ElasticNet`)
- Random Forest (`RandomForestClassifier`)
- Light Gradient Boosting Machine (`LightGBM`)

- Extreme Gradient Boosting (`XGBoost`)
- TabPFN (pretrained foundation model)
- Decision Tree (CART)

For each model, a three-fold stratified cross-validation strategy was used on the training set for hyperparameter optimisation using Bayesian search via `scikit-optimize`. Performance was evaluated using AUROC, average precision (AP), F1 score, precision, and recall. The final model was refit on the full training set before evaluating on the test set.

#### 2.3 Hyperparameter Search Spaces and Final Selections

**ElasticNet:**

- `penalty` (L1/L2 mixing ratio): [0.1, 0.5, 0.9]
- `alpha` (regularisation strength): log-uniform $[10^{-4}, 10^{1}]$

**Random Forest:**

- `n_estimators`: 100, 250, **500**
- `max_depth`: None, 5, 10, 20
- `min_samples_leaf`: 1, 2, **5**
- `class_weight`: 'balanced'

**LightGBM:**

- `num_leaves`: 15, 31, 63
- `min_data_in_leaf`: 5, 10, 20
- `learning_rate`: 0.1, 0.01, **0.005**
- `n_estimators`: **500**

**XGBoost:**

- `learning_rate`: 0.01, 0.05, **0.1**
- `max_depth`: 3, **5**, 7
- `subsample`: 0.5, 0.8, 1.0
- `n_estimators`: 100, **300**

**TabPFN:** used without tuning; we employed the pretrained model provided by the authors on the OpenML default setting with 10,000 budgeted inference steps.

**Decision Tree:**

- `max_depth`: 3, 5, 7, None
- `criterion`: 'gini', 'entropy'

##### 2.4 Model Evaluation and Metrics

For each bootstrap iteration, models were trained using the tuned parameters and evaluated on the held-out test set. We report the mean and standard deviation across the 10 resamples for the following metrics:

- Area Under the Receiver Operating Characteristic Curve (AUROC)

$$\text{AUROC} = \int_0^1 TPR(FPR^{-1}(x))dx$$

- Average Precision (AP)

$$\text{AP} = \sum_n (R_n - R_{n-1})P_n$$

  where $P_n$ and $R_n$ are precision and recall at threshold $n$.

- F1 Score

$$\text{F1} = 2 \cdot \frac{Precision \cdot Recall}{Precision + Recall}$$

- Precision and Recall

$$\text{Precision} = \frac{TP}{TP + FP}, \quad \text{Recall} = \frac{TP}{TP + FN}$$

To account for the small number of positive cases ($N = 13$ across the full cohort), we used stratified bootstrapping and reported confidence intervals. Feature importance was derived directly from impurity-based Random Forest scores averaged across bootstrap splits.

#### 3 Foundation Model Setup and Prompting

To evaluate the zero-shot predictive capability of foundation models for cardiovascular disease (CVD) risk, we designed controlled prompting procedures for NormWear and PaPaGei, each operating in inference-only mode on electrocardiogram (ECG) and photoplethysmography (PPG) signals, respectively.

##### 3.1 NormWear Prompt Design

For NormWear experiments, we used the `NormWearZeroShot` PyTorch implementation, loading pretrained weights and model-specific tokenisers as described in the original release. NormWear accepts paired ECG signals and text prompts, performing multimodal alignment via contrastive inference. We used the following prompt setup:

- **Query:** `"This is a 3-lead ECG from a patient.  What does it indicate?"`

- **Options:**
  - `"The patient has no signs of cardiovascular disease."`
  - `"The patient shows signs of cardiovascular disease or stress."`

Each 3-lead ECG was segmented into 2-second windows (128 samples at 64 Hz) and up to 20 high-quality windows per patient were selected using signal quality control filters. Per-window predictions were generated using similarity scores between the ECG embeddings and text embeddings. These predictions were aggregated at the patient level using majority voting and averaged probabilities. No task-specific fine-tuning was performed, and the model operated fully in zero-shot mode.

#### 3.2 PaPaGei Prompt-Free Inference

In contrast, PaPaGei is a general-purpose encoder model producing dense time-series embeddings without explicit prompting. We employed the `ResNet1DMoE` architecture with the following configuration:

- `base_filters = 32`, `kernel_size = 3`, `stride = 2`
- `n_block = 18`, `n_experts = 3`, `embedding dimension = 512`

Each 10-second PPG window (resampled to 100 Hz) was denoised, z-score normalised, and filtered using Vital-SQI criteria to remove poor-quality segments. Embeddings were extracted from the penultimate layer of the pretrained model. For downstream classification:

1. Window-level embeddings were aggregated per patient via mean pooling.
2. Dimensionality reduction was performed via principal component analysis (PCA, top 15 components).
3. A Random Forest classifier was trained on patient-level embeddings using 10 bootstrapped train-test splits.
4. Model calibration was performed using isotonic regression on the training set.

No fine-tuning or additional supervision was applied to PaPaGei during this process, consistent with the zero-shot foundation model paradigm.

#### 3.3 Inference Code Snippets

**NormWear prompting setup:**

```
task = ["This is a 3-lead ECG from a patient. What does it indicate?"]
options = [
    "The patient has no signs of cardiovascular disease.",
    "The patient shows signs of cardiovascular disease or stress."
]
txt_embed = model.txt_encode(task + options)
query_embed, option_embed = txt_embed[:1], txt_embed[1:]
signal_embed = model.signal_encode(ecg_tensor_batch, query_embed)
probs = model.inference(signal_embed, option_embed)
```

**PaPaGei embedding extraction:**

```
model = ResNet1DMoE(**model_config).to(device)
model.load_state_dict(torch.load("papagei_s.pt"))
model.eval()
embedding = model(signal_tensor)  # Shape: [batch_size, 512]
```

A complete list of preprocessing steps, model parameters, and inference scripts is available at our code repository [link redacted for peer review].

#### References
